# Immune response to 2-dose BNT162b2 vaccination and risk of SARS-CoV-2 breakthrough infection: The Shieldvacc-2 study

**DOI:** 10.1101/2022.04.19.22273872

**Authors:** Lisa Seekircher, Zoltán Bánki, Janine Kimpel, Annika Rössler, Helena Schäfer, Barbara Falkensammer, David Bante, Lukas Forer, Sebastian Schönherr, Teresa Harthaller, Magdalena Sacher, Cornelia Ower, Lena Tschiderer, Hanno Ulmer, Florian Krammer, Dorothee von Laer, Wegene Borena, Peter Willeit

## Abstract

It is uncertain to which extent antibody and T-cell responses after vaccination against SARS-CoV-2 are associated with reduced risk of breakthrough infection and whether their measurement enhances risk prediction. We conducted a phase-4 open-label clinical trial in the pre-omicron era, enrolling 2,760 individuals aged ≥16 years 35±8 days after having received the second dose of BNT162b2 (baseline 15-21 May 2021). Over a median 5.9-month of follow-up, we identified incident SARS-CoV-2 breakthrough infections using weekly antigen tests, a confirmatory PCR test, and/or serological evidence for incident infection. We quantified relative risks adjusted for age, sex, and prior SARS-CoV-2 infection for different immunological parameters and assessed improvements in risk discrimination. In contrast to the T-cell response, higher plasma levels of binding antibodies and antibodies in a surrogate neutralization assay were associated with reduced risk of breakthrough infection. Furthermore, assessment of anti-spike IgG levels enhanced prediction of breakthrough infection and may therefore be a suitable measurable correlate of protection in practice.

## Introduction

Measurable correlates of protection that help assess to which extent a person is protected from severe acute respiratory syndrome coronavirus 2 (SARS-CoV-2) infection after vaccination (so-called ‘breakthrough infection’) are useful for the individual, but also to estimate the degree of a population’s protection. Such candidate biomarkers are plasma levels of antibodies directed against the SARS-CoV-2 spike and nucleocapsid proteins, SARS-CoV-2 neutralizing antibody levels, and markers of the cellular response to vaccination. Several prior studies have suggested inverse associations between these markers and risk of breakthrough infection.^1–10^ However, there is uncertainty about the shapes of associations (i.e., linear or non-linear, presence of threshold), few prior studies dispose of concurrent measurements of a broad range of immunological parameters, including the cellular response, and the bulk of studies have been conducted in select population subgroups. Furthermore, it is unclear whether measurement of these immunological parameters enhances prediction of breakthrough infection risk over and beyond more readily available information such as age and sex.

To provide clarity, we conducted a phase-4 open-label clinical trial among 2,760 individuals vaccinated with two doses of BNT162b2, who live in the district of Schwaz in the Federal State of Tyrol, Austria. Our aims were two-fold. First, to estimate relative risks of incident SARS-CoV-2 infection according to levels of several humoral and cellular immune parameters. Second, to quantify the predictive value of binding and neutralizing antibodies for incident SARS-CoV-2 infection.

## Results

### Characteristics of participants

Study enrolment took place between 15 and 21 May 2021. Of 3,268 eligible individuals, we excluded 12 without a baseline blood sample and 496 individuals without follow-up information (i.e., no antigen test during the study and no test result of Ig antibodies targeting the nucleocapsid protein [anti-N Ig] at end of the study), leaving a total of 2,760 individuals for further analysis. **Table 1** summarizes the baseline characteristics of these individuals, overall and according to incident SARS-CoV-2 infection. Overall, median age was 47.4 years (standard deviation [SD] 14.5) and 60.9% were female. 25.8% had a prior SARS-CoV-2 infection, which had occurred a median of 6.1 months before the study’s baseline (interquartile range [IQR] 5.0-6.6). Median time since vaccination was 67 days (IQR 65-68) for dose 1 and 39 days (37-40) for dose 2. Frequencies of reported complaints that occurred within one week post-vaccination are shown in **Supplementary Fig. 1**. Vaccinations were generally well tolerated. The most common complaints were pain at the injection site (dose 1: 67.6%, dose 2: 53.2%), fatigue (dose 1: 21.8%, dose 2: 40.1%), and headache (dose 1: 11.9%, dose 2: 23.5%).

**Table 1:**
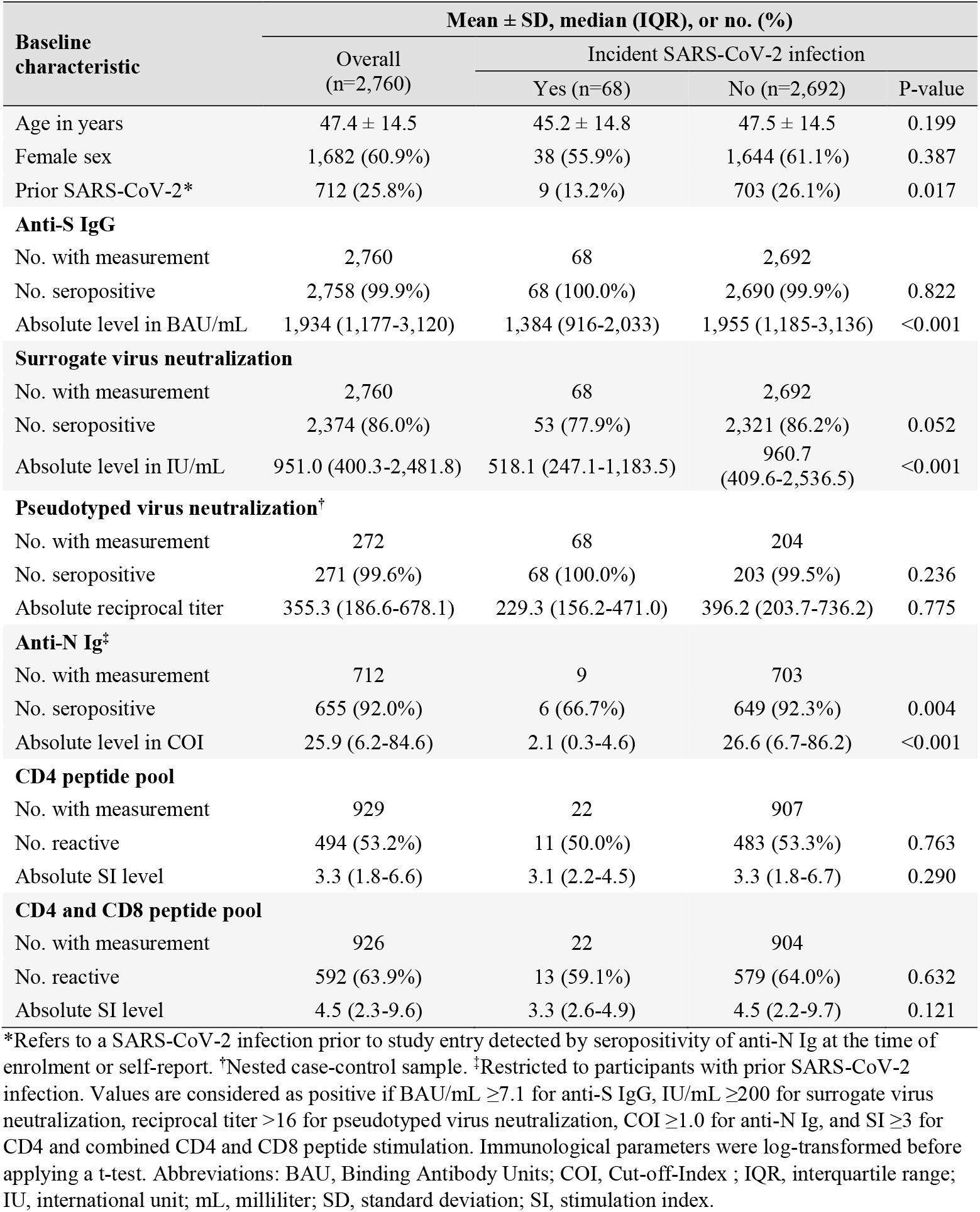
Baseline characteristics of the entire study population and separately for participants with and without incident SARS-CoV-2 infection during follow-up.

Baseline immunological parameters are also described in **Table 1**. Except for two immunosuppressed participants, all individuals were seropositive for IgG antibodies targeting the receptor binding domain (RBD) of the spike protein (anti-S IgG) and 86.0% had seropositive titers of neutralizing antibodies in a surrogate SARS-CoV-2 virus neutralization test (sVNT). Among the 712 participants with prior SARS-CoV-2 infection, 655 were seropositive for anti-N Ig (92.0%). Titers of anti-S IgG, neutralizing antibodies in the sVNT, and anti-N Ig were all significantly lower in participants who had a SARS-CoV-2 infection during follow-up as compared to those who never acquired the infection during the follow-up (all P<0.001). Titers of neutralizing antibodies in a pseudotyped SARS-CoV-2 virus neutralization test (pVNT) were measured in a subset of 68 cases and 204 individual-matched controls with 271 seropositive samples and a median reciprocal titer level of 355.3 (IQR 186.6-678.1). Among the participants in whom cellular response was analyzed, 53.2% showed reactivity for CD4 peptide pool and 63.9% were reactive for SARS-CoV-2-derived combined CD4 and CD8 peptides. Median stimulation index (SI) levels of CD4 and combined CD4 and CD8 peptides were 3.3 (IQR 1.8-6.6) and 4.5 (2.3-9.6), respectively.

Scatter plots and correlations of different baseline immunological parameters are depicted in **Fig. 1**. There was a strong positive correlation of levels of anti-S IgG with titers of neutralizing antibodies in the sVNT (r = 0.79 [95% CI 0.77-0.80]), and the pVNT (0.82 [0.78-0.86]). In comparison, these parameters correlated more weakly with anti-N Ig. Also, correlations between humoral and cellular immune parameters were low, ranging from 0.24 to 0.34, whereby responses to CD4 and combined CD4 and CD8 peptide pools were very highly correlated with each other (0.92 [0.91-0.93]).

**Fig. 1:**
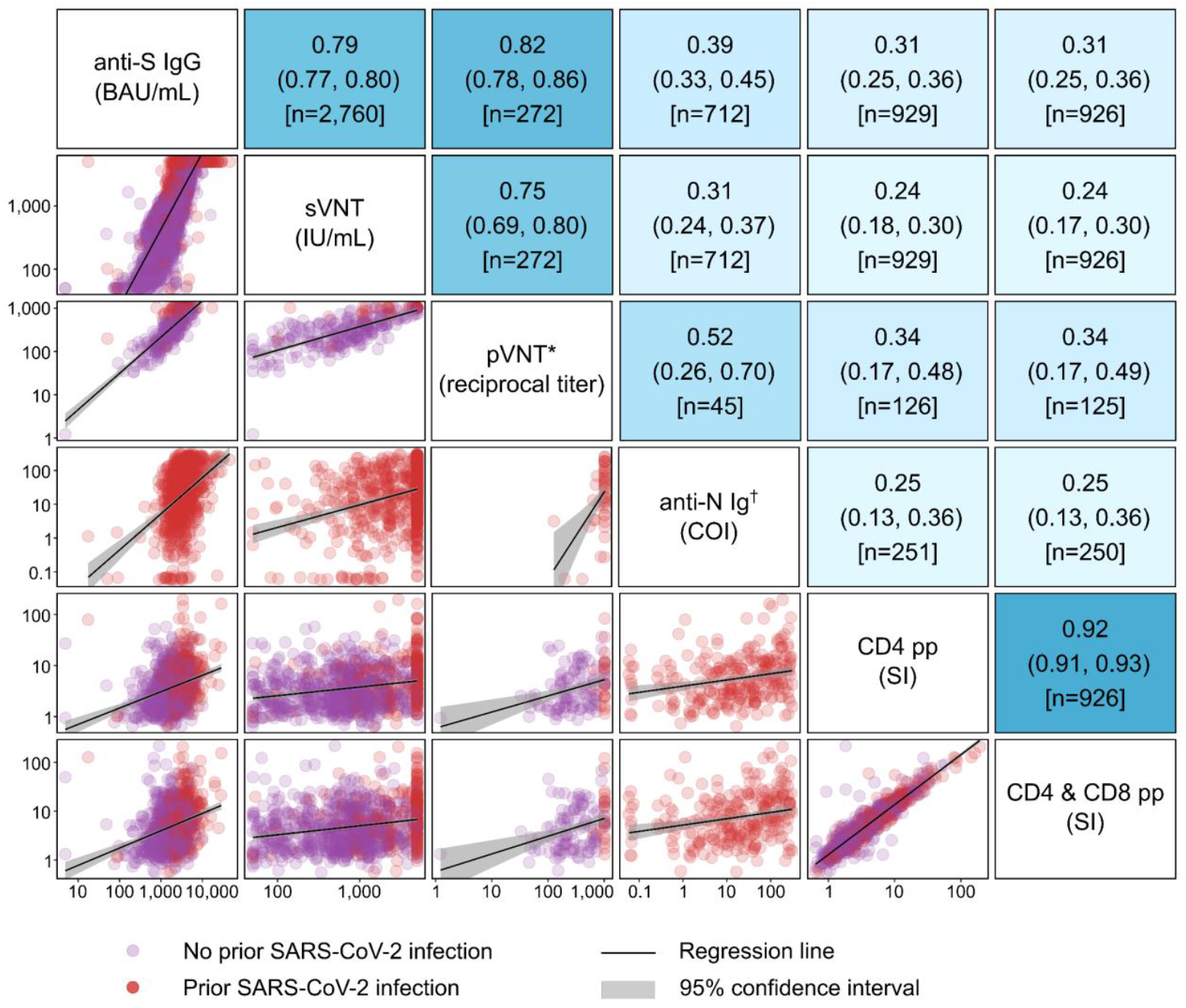
Correlation coefficients and scatter plots of the baseline levels of the immunological parameters assessed in the study. *The analyses of pVNT values were restricted to the nested case-control sample. ^†^The analyses of anti-N Ig values were restricted to participants with prior SARS-CoV-2 infection. For all analyses log-transformed immunological parameters were used. The upper part of the matrix indicates unadjusted Pearson correlation coefficients with 95% confidence intervals and number of participants. Areas are shadowed according to the magnitude of the point estimates of Pearson correlation coefficients, i.e., the darker the closer to one, the lighter the closer to zero. The lower part of the matrix depicts scatter plots of different immunological parameters, with both axes presented on a log scale. Abbreviations: BAU, Binding Antibody Units; COI, Cut-off-Index ; IU, international unit; mL, milliliter; pp, peptide pool; pVNT, pseudotyped virus neutralization test; SI, stimulation index; sVNT, surrogate virus neutralization test.

### Incident SARS-CoV-2 infections and their association with immunological parameters

**Supplementary Fig. 2** shows the cumulative incidence curve of SARS-CoV-2 breakthrough infection among Shieldvacc-2 participants and the population in the district of Schwaz, which both sharply increased in the last third of follow-up (i.e., autumn 2021). Over a median follow-up of 5.9 months (IQR 5.8-5.9) corresponding to 14,995 person-days at risk, we recorded 68 events of a SARS-CoV-2 breakthrough infection. Breakthrough infections happened between 1 August and 15 November 2021 and consequently the majority should be caused by the Delta variant (B.1.617.2) as this was the dominant variant during this time. Fifty-three (77.9%) of the breakthrough infections were symptomatic. The frequencies of reported symptoms are shown in **Supplementary Fig. 3**. The most common symptoms were cough (57.4%), loss of taste or smell (45.6%), muscle or limb pain (44.1%), and fever or chills (36.8%). One case of SARS-CoV-2 infection required hospitalization; none were fatal. No study participant died from other causes.

The relative risks for breakthrough infection per two-fold higher level were 0.72 (95% confidence interval [CI]: 0.60-0.86; P<0.001) for anti-S IgG and 0.80 (0.69-0.92; P=0.002) for titers of neutralizing antibodies in the sVNT (**Fig. 2**). In contrast, we observed no significant associations for neutralizing antibodies in the pVNT (odds ratio: 0.79 [95% CI: 0.62-1.01; P=0.063]) and of cellular immunity in response of neither CD4 nor the combination of CD4 and CD8 peptide pools with incident SARS-CoV-2 infection, yielding relative risks of 0.84 (0.58-1.21; P=0.343) and 0.77 (0.54-1.08; P=0.129), respectively.

**Fig. 2:**
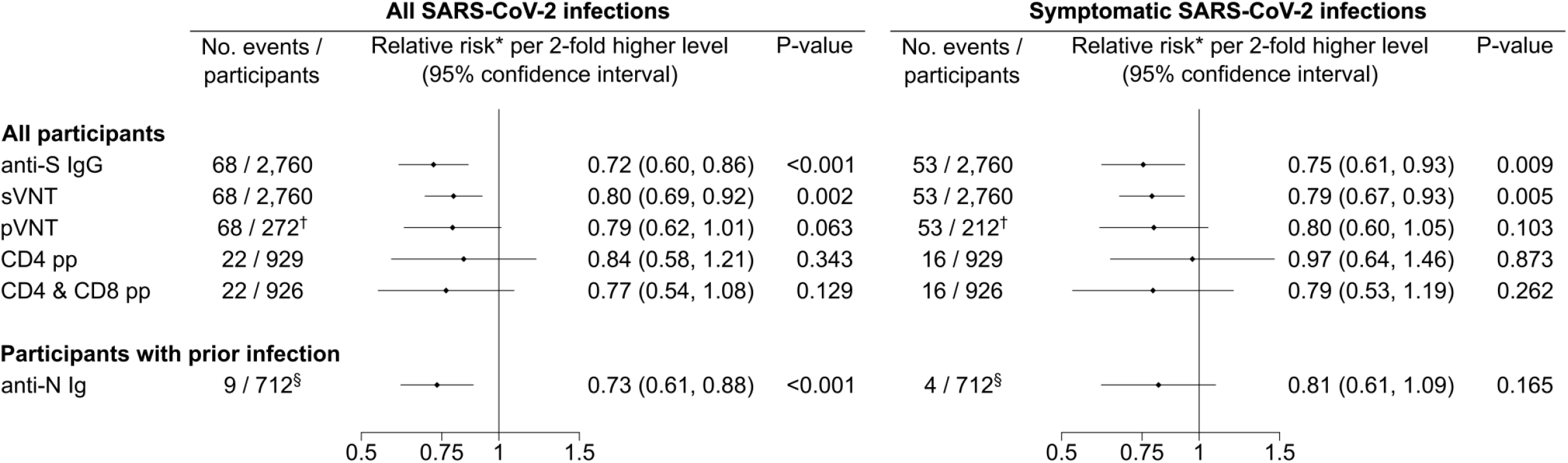
Relative risk of SARS-CoV-2 infection according to baseline levels of immunological parameters. *Relative risks were adjusted for age, sex, and prior SARS-CoV-2 infection. ^†^pVNT was measured in a subset of 68 cases and 204 individual-matched controls. ^‡^pVNT was measured in a subset of 53 symptomatic cases and 159 individual-matched controls. ^§^The analysis of anti-N Ig was restricted to participants with prior SARS-CoV-2 infection. Prior SARS-CoV-2 infection was based on self-report or seropositivity of anti-N Ig at the time of enrolment. **Supplementary Table 1** provides additional information on participants with prior SARS-CoV-2 infection and with incident SARS-CoV-2 breakthrough infection. Symptomatic SARS-CoV-2 infection was defined as having one or more symptoms including fever or chills, cough, breathing difficulties, muscle or limb pain, lost sense of smell or taste, sore throat, diarrhea, or vomiting. Cox regression was applied for anti-S IgG, sVNT, CD4 pp, CD4 & CD8 pp, and anti-N Ig and conditional logistic regression for pVNT. Immunological parameters entered as log2-transformed continuous terms. Abbreviations: pp, peptide pool; pVNT, pseudotyped SARS-CoV-2 virus neutralization test; sVNT, surrogate SARS-CoV-2 virus neutralization test.

Among the 712 participants with a SARS-CoV-2 infection prior to the study, 9 (1.3%) were re-infected during follow-up (for details, see **Supplementary Table 1**). The relative risk of breakthrough infection for anti-N Ig in the group with prior infection was 0.73 (0.61-0.88; P<0.001).

Secondary analyses restricted to symptomatic SARS-CoV-2 infections yielded broadly similar results (**Fig. 2**). Additional secondary analyses quantified relative risks across different categories of levels of immunological parameters (**Fig.3**). In all analyses, P-values for trend were 0.001 or lower and shapes of associations were log-linear, with no evidence of any thresholds that would clearly delineate population groups at high vs. low risk. For example, we observed a significantly lower relative risk for individuals with anti-S IgG levels ≥3,000 Binding Antibody Units per milliliter (BAU/mL) compared to those with <500 BAU/mL (hazard ratio: 0.20 [0.06-0.64]; P=0.007) and for individuals with anti-N Ig levels ≥25 Cut-off-Index (COI) compared to those seronegative with <1 COI (hazard ratio: 0.09 [0.01-0.67]; P=0.019). Cumulative incidence plots according to categories of immunological parameters are depicted in **Supplementary Fig. 4**.

**Fig. 3:**
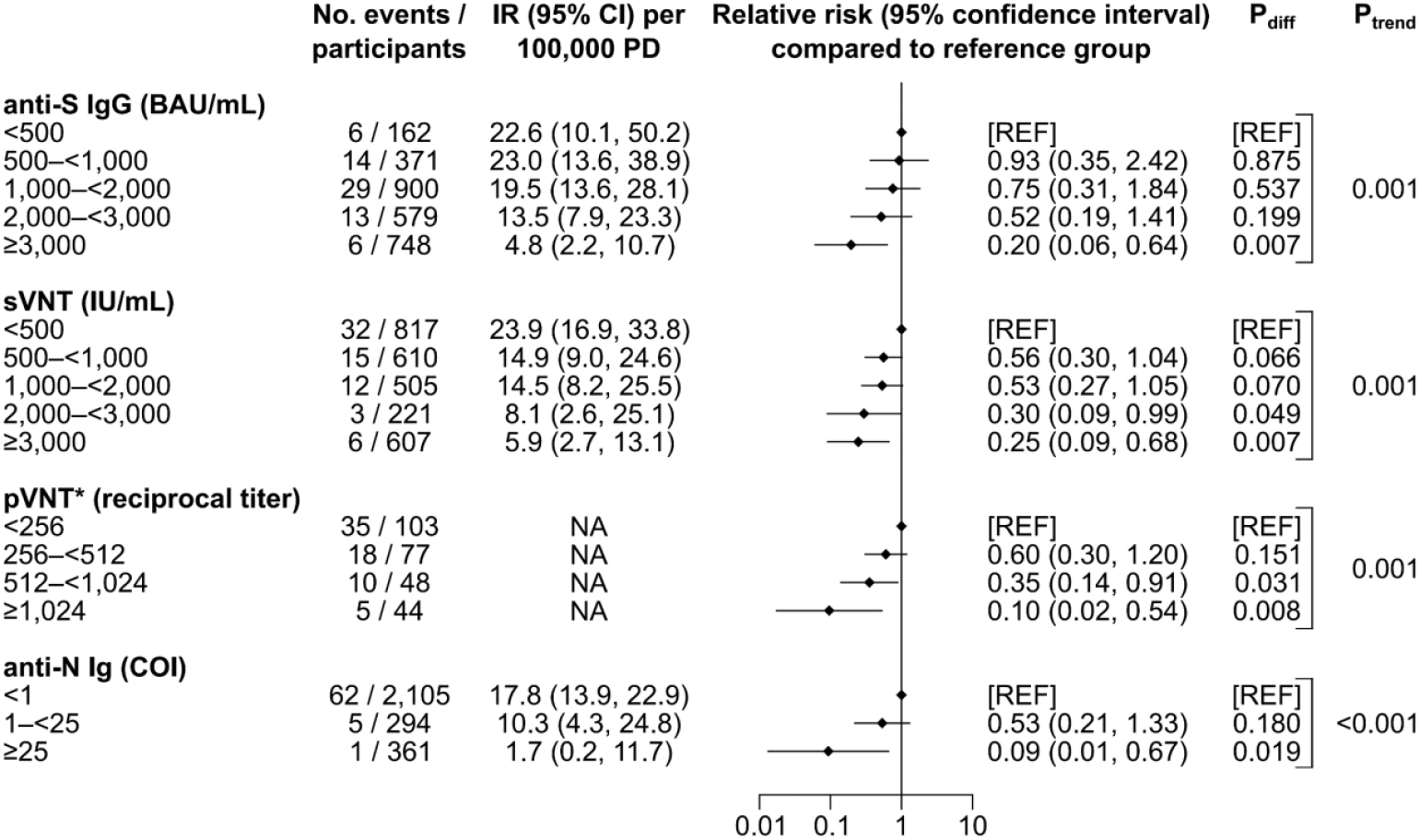
Relative risk of SARS-CoV-2 infection across categories of baseline levels of immunological parameters. *pVNT was measured in a subset of 68 cases and 204 individual-matched controls. Cox regression was applied for anti-S IgG, sVNT, and anti-N Ig and conditional logistic regression for pVNT. For anti-N Ig, the regression model was adjusted for age and sex (and not for prior SARS-CoV-2 infection due to collinearity) and for anti-S IgG, sVNT, and pVNT additionally for prior SARS-CoV-2 detected by seropositivity of anti-N Ig at the time of enrolment or self-report. P_trend_ indicates the p-value of the likelihood ratio test comparing regression models including categories of antibody levels as a continuous variable and without antibody information. **Supplementary Table 1** provides additional information on participants with prior SARS-CoV-2 infection and with incident SARS-CoV-2 breakthrough infection. Abbreviations: BAU, Binding Antibody Units; CI, confidence interval; COI, Cut-Off-Index; IR, incidence rate; IU, international unit; NA, not available; mL, milliliter; PD, person-days; pVNT, pseudotyped SARS-CoV-2 virus neutralization test; sVNT, surrogate SARS-CoV-2 virus neutralization test.

### Predictive value of assessing immunological parameters for incident SARS-CoV-2 infection

To assess the incremental value of immunological parameters for predicting SARS-CoV-2 infection, we quantified improvements in the C-index when adding their measurements to a common base model (**Fig. 4**). The base model incorporated information on age and sex and had a C-index of 0.562 (95% CI: 0.494-0.631). The separate addition of immunological parameters provided significant improvements in the C-index by 0.085 (0.027-0.143; P=0.004) for anti-S IgG, 0.080 (0.013-0.147; P=0.020) for neutralizing antibodies in the sVNT, 0.054 (0.002, 0.106; P=0.040) for anti-N Ig, and 0.088 (0.026-0.151; P=0.006) for the combination of anti-S IgG and anti-N Ig. The latter did not yield a significantly higher C-index than measurement of anti-S IgG alone (0.004 [-0.030, 0.038]; P=0.824). As a benchmark, information on self-reported prior infection provided a C-index change of (0.015 [-0.037, 0.067]; P=0.575), when added to the base model.

**Fig. 4:**
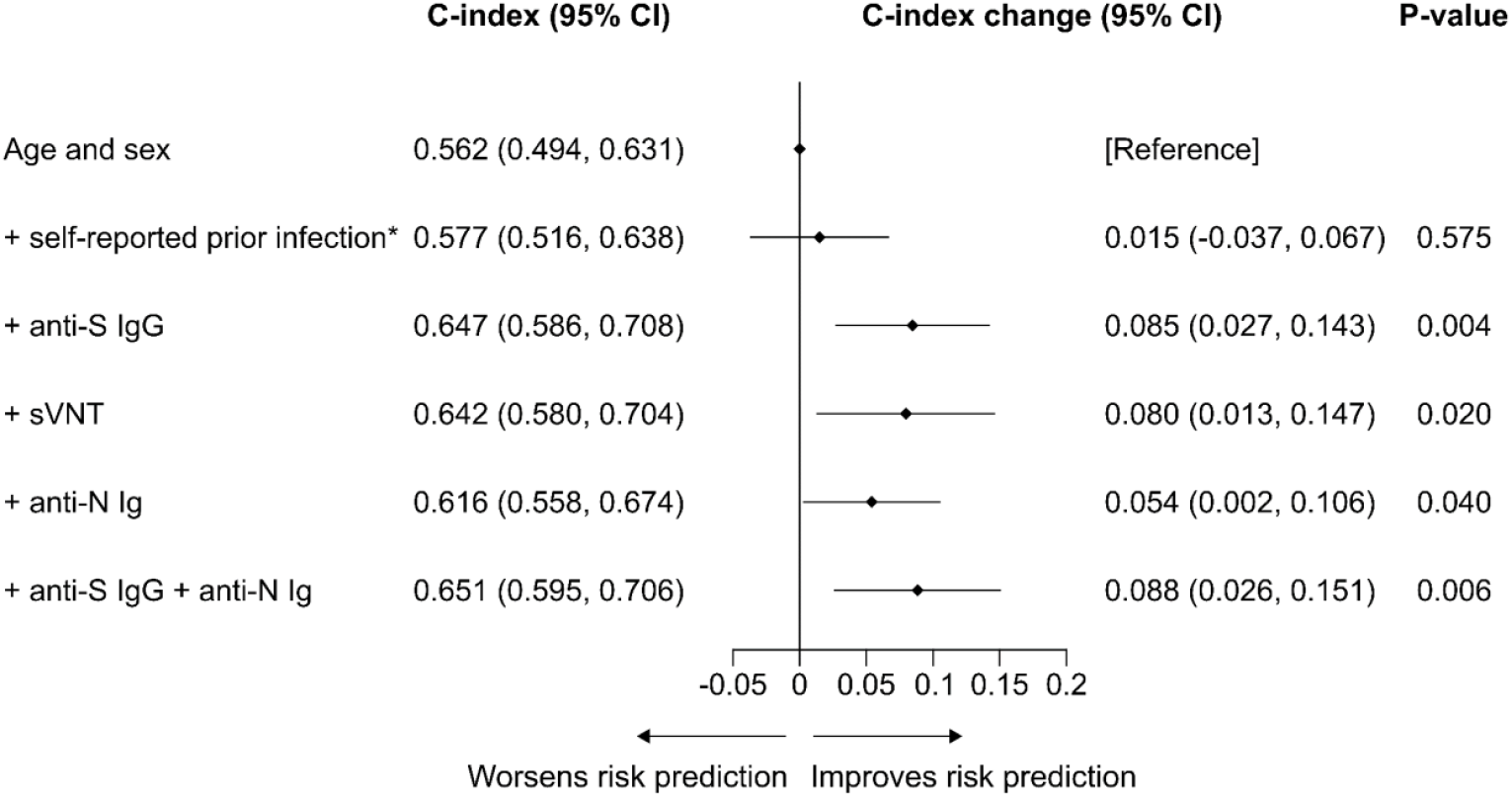
Improvement in prediction of incident SARS-CoV-2 infection upon additional assessment of information on anti-SARS antibodies and prior SARS-CoV-2 infection. *Refers to a SARS-CoV-2 infection prior to study entry detected by self-report. Participants with complete data on all variables are included in analyses (2,760 participants; 68 incident SARS-CoV-2 events). We quantified improvements in the C-index when adding them to a model containing information on age and sex. Immunological parameters entered as log2-transformed continuous terms. Abbreviations: CI, confidence interval; sVNT, surrogate SARS-CoV-2 virus neutralization test.

## Discussion

In the present study involving 2,760 participants aged 16 years or older, we evaluated humoral and cellular immunological parameters after 2-dose BNT162b2 vaccination as potential correlates of protection against SARS-CoV-2 infection over a six-month follow-up period. We observed strong inverse log-linear associations with risk of incident SARS-CoV-2 breakthrough infections for a number of immunological parameters, i.e., anti-S IgG, titers of neutralizing antibodies in a sVNT, and – in people that had a SARS-CoV-2 infection before study inclusion – levels of anti-N Ig antibodies. In contrast, there was no significant association for levels of neutralizing antibodies in a pVNT and of cellular immune response to vaccination with breakthrough infection risk. Finally, we provide novel data on the usefulness of anti-S IgG in the prediction of breakthrough infection, showing that assessment of anti-S IgG provided a substantial improvement in risk discrimination over and beyond a model containing information on age and sex.

Our findings corroborate previous data from clinical trials and observational studies showing inverse relationships between humoral immune responses to vaccination and subsequent risk of breakthrough infection. The COV002 trial – a phase-2/3 trial of the ChAdOx1 vaccine – measured anti-S IgG, anti-RBD IgG, and titers of neutralizing antibodies in a pseudotyped and a live-virus neutralization assay 28 days after receipt of the second dose and found that higher levels were linked to a significantly reduced risk of symptomatic infection over a ∼3-month follow-up.^1^ Similarly, the COVE trial – a phase-3 trial of mRNA-1273 – reported after an average follow-up period of 2.7 months that 10-fold higher levels of anti-S IgG, anti-RBD IgG, and neutralizing antibodies in a pVNT measured 28 days after second dose were associated with hazard ratios for breakthrough infection of 0.66 (0.50-0.88), 0.57 (0.40-0.82), and 0.42 (0.27-0.65), respectively.^2^ In comparison to these trials, effect sizes for anti-S IgG in our study were stronger and were robust in analysis of the pre-specified primary outcome of “any infections” as well as in a sensitivity analysis restricted to symptomatic infections.

Our results are also in agreement with previous observational studies that were conducted in vaccinated community samples,^3–5^ health-care workers,^6^ patients with autoimmune rheumatic diseases,^7^ and patients undergoing dialysis^8–10^. However, while the majority of studies compared risk across categories (e.g., dichotomizing the study population at arbitrary cut-offs for anti-S IgG levels), our study revealed associations compatible with a log-linear shape, thereby suggesting the higher the level of immune response the lower the risk of breakthrough infection without evidence for a threshold or saturation effect.

For T-cell responses, we did not detect measurable differences in post-vaccination levels between people with or without breakthrough infection, which is in concordance with the findings of studies among BNT162b2-vaccinated individuals from Italy^11^ and Singapore^12^. This is may be explained by the main function of T-cells, which is to facilitate early viral clearance^13^, and hence circumvent severe clinical course, rather than prevent a primary infection per se. The shorter incubation time of the Delta variant (2-3 days)^14^ may also limit the potential of T-cells to avert symptomatic disease, while T-cells have more time to respond to protect from severe disease. The fact that the majority of breakthrough infections were rather of mild nature may support this notion making it impossible to detect any appreciable difference in T-cell response between groups. That the major mechanism of protection from acquiring the infection comes from neutralizing antibodies is further supported by previous data portraying non-significant effect of vaccine-induced antibodies in preventing infection with variants – for example the Omicron variant (B.1.1.529) – highly mutated at the binding sites of neutralizing antibodies.^15,16^ Our study ended before the emergence of the Omicron variant in the region, making it impossible to comment on any change in the breakthrough infection pattern.

In our study, we also evaluated the added value of assessing anti-SARS-CoV-2 antibodies for predicting a person’s risk of developing SARS-CoV-2 despite vaccination. Based on our findings in the risk discrimination analysis and on lower complexity and cost, anti-S IgG appears to be the most suitable measurable correlate of protection in practice, yielding a large improvement in the C-index by 0.085 (95% CI: 0.027-0.143; P=0.004). Anti-N Ig antibody measurement showed no incremental predictive value when jointly measured with anti-S IgG. No prior study has investigated the usefulness of these immunological parameters in SARS-CoV-2 risk prediction and, therefore, our study provides entirely novel evidence on this topic.

In another set of analyses, we examined the cross-sectional correlations of different immunological parameters elicited by vaccination. Together with evidence from other studies^2,14,17–22^, the strong correlation we observed between anti-S IgG and neutralizing antibodies indicates a high potential of anti-S IgG to quantitatively reflect neutralizing capacities for variants before the emergence of Omicron. In participants with a prior SARS-CoV-2 infection, the only moderate correlation of anti-N Ig with other parameters can be explained by the stronger waning of anti-N Ig over time after SARS-CoV-2 infection.^23^ Furthermore, the anti-S IgG response is steered by previous infection and vaccination, whereas anti-N Ig response is purely post-infection which potentially distorts the correlation. This notion is supported by previous data from pre-SARS-CoV-2 vaccine era that showed a considerable degree of correlation between anti-N and anti-S IgG antibodies generated post-infection.^24^ Our finding of poor correlations between antibodies and T-cell responses is consistent with some other studies showing no or low correlations between humoral and cellular immune parameters in vaccinated^25–30^ and convalescent^31–33^ individuals.

Our study has several strengths. It has a prospective study design, is adequately sized, covers a ∼6 months follow-up post second dose, and compared immunological parameters for humoral and cellular immunity measured with validated assays. To the best of our knowledge, our study is the first to analyze associations between T-cells and incident SARS-CoV-2 infection using time-to-event analysis. Furthermore, all incident SARS-CoV-2 infections and related symptoms were validated rigorously in structured telephone interviews. Our study also has limitations. First, cellular immune parameters were available only for a (random) subgroup of participants, thereby limiting statistical power. Second, the QuantiFERON SARS-CoV-2 assay was limited to measure the Interferon-Gamma (IFN-γ) production after stimulation with CD4 and combined CD4 and CD8 peptide pools, therefore a detailed characterization of T-cell response with respect of the source of IFN-γ (CD4 or CD8 T-cells), phenotypical and further functional analysis of T-cells is missing. Third, the proportion of participants with a prior SARS-CoV-2 infection was relatively high (25.8%) and is likely due to high interest to participate in the study. Ascertainment of prior infection was of high quality as it was performed by trained study staff and was confirmed by a positive anti-N Ig measurement in 92% of cases. Fourth, we conducted our study during a phase in which Delta was the predominant variant of concern and associations of immunological parameters with Omicron may be weaker. Finally, these analyses have been conducted on samples taken after two doses of BNT162b2 and might not apply to other coronavirus disease 2019 (COVID-19) vaccines or to people that received a third doses of BNT162b2.

In conclusion, in contrast to the T-cell response, higher plasma levels of binding and neutralizing antibodies in a surrogate neutralization assay were associated with reduced risk of breakthrough infection. Assessment of anti-S IgG levels enhances prediction of incident SARS-CoV-2 infection and may therefore be a suitable measurable correlate of protection in practice.

## Methods

### Study design and participants

The Shieldvacc-2 study is a phase-4 open-label clinical trial conducted at two centers (Jenbach, Zell am Ziller) in the district of Schwaz, Tyrol, Austria. Individuals were eligible for inclusion if they (i) were aged 16 years or older; (ii) had received two 30 µg doses of BNT162b2 delivered by intramuscular injection, with the second dose having been administered 35±8 days prior to enrolment into the study; (iii) understood and agreed to comply with the study procedures; and (iv) were willing to be contacted by telephone or to complete a diary throughout the study in an online participant portal. Exclusion criteria were: (i) prior administration of an investigational coronavirus (SARS-CoV, middle east respiratory syndrome coronavirus) vaccine or current/planned simultaneous participation in another interventional study to either prevent or treat COVID-19; (ii) a contraindication to blood draws (e.g., bleeding disorders); and (iii) participation in an interventional clinical study within the 30 days prior to study enrolment. Eligible persons were invited by public calls in the radio and newspaper to participate in the study.

At the study baseline between 15 and 21 May 2021, participants were asked to complete a questionnaire covering detailed information on sociodemographic characteristics, prior SARS-CoV-2 infection, and SARS-CoV-2 vaccination. In addition, blood samples of up to 18 mL were drawn to allow testing of the participants’ humoral and cellular immune responses to vaccination. Six months after the study baseline between 11 and 18 November 2021, participants were again invited for blood draws.

Written informed consent was provided by study participants or – if appropriate – by the individual’s legal representative or custodial. The study was approved by the ethics committee of the Medical University of Innsbruck (no. 1168/2021) and has been registered at the European Union Drug Regulating Authorities Clinical Trials Database (EudraCT number: 2021-002030-16).

### Laboratory measurements

Details on laboratory methods are provided in the **Supplementary Material**. In brief, to assess antibody responses, plasma samples were collected in S-Monovette tubes (Sarstedt, Nümbrecht, Germany) containing ethylenediaminetetraacetic acid anticoagulant (EDTA KE/9 mL) and were analyzed with three distinct commercially available serological tests: (i) the Abbott SARS-CoV-2 IgG II Quant chemiluminescent microparticle immunoassay on the Alinity i instrument (Abbott Ireland, Sligo, Ireland) to measure anti-S IgG; (ii) the Roche Elecsys Anti-SARS-CoV-2 electrochemiluminescent immunoassay on the Cobas e411 analyzer (Roche, Mannheim, Germany) to measure anti-N Ig; and (iii) the TECO SARS-CoV-2 Neutralization Antibody enzyme-linked immunosorbent assay on the SERION Immunomat (TECOmedical, Sissach, Switzerland) to measure inhibitory effects of neutralizing antibodies blocking the interaction of angiotensin-converting enzyme 2 and RBD of SARS-CoV-2 spike protein. In addition to these commercially available serological tests, in a sample of 68 cases and 204 controls individually matched by age, sex, and prior SARS-CoV-2 infection, we determined 50% neutralizing antibody titers against the ancestral (Wuhan-1) spike using a vesicular stomatitis virus pseudovirus assay^34,35^ with reciprocal titers >16 being regarded as positive.

Furthermore, to evaluate cellular immune responses, in a random subgroup of 929 participants, additional blood samples were collected in S-Monovette tubes (Sarstedt, Nümbrecht, Germany) containing lithium-heparin anticoagulant (Li-Heparin LH/9 mL). SARS-CoV-2-specific T-cell response was measured by Qiagen QuantiFERON SARS-CoV-2 RUO IFN-γ release assay (Qiagen, Hilden, Germany) in response to CD4 and combined CD4 and CD8 peptide pools derived from SARS-CoV-2 spike antigen (S1 S2 RDB). The ratios of IFN-γ values from SARS-CoV-2 specific stimulation and the unstimulated control (Nil) was determined as the SI. We considered samples with SI values <2 as negative, 2≤ SI <3 as weakly reactive, and values ≥3 as reactive. All samples were processed centrally at the laboratory of the Institute of Virology of the Medical University of Innsbruck, Austria.

### Outcome definition and ascertainment

The primary outcome was defined as the occurrence of a SARS-CoV-2 breakthrough infection during a follow-up period of six months, identified by a positive PCR test (n=50), a seroconversion of anti-N Ig from baseline to follow-up (n=14), or a threefold increase of a positive anti-N Ig level during the study period (n=4). To preclude underascertainment of asymptomatic or pauci-symptomatic events, participants were asked to undergo SARS-CoV-2 antigen testing every 7 (±3) days throughout the course of the study and to record test results and related symptoms via the online participant portal. A secondary analysis focused on symptomatic SARS-CoV-2 infections defined as in the BNT162b2 phase-2/3 trial as having one or more symptoms of fever or chills, cough, breathing difficulties, muscle or limb pain, lost sense of smell or taste, sore throat, diarrhea, or vomiting.^36^ All recorded SARS-CoV-2 infections, including dates of infections, symptoms, and clinical course, underwent rigorous validation in structured telephone interviews. For events detected through serological tests only, the date of infection was estimated using the dates of plausible contagions (e.g., symptoms or close contact with infected individuals) (n=12) or otherwise using the median date of all SARS-CoV-2 events that were recorded in the study (n=6). Prior SARS-CoV-2 infection was based on self-report or seropositivity of anti-N Ig at the time of enrolment.

### Statistical analysis

Because the distributions of immunological parameters were skewed, we log-transformed their values for all analyses. We used t-tests for continuous variables and χ^2^-tests for categorical variables to compare baseline characteristics of participants with and without incident SARS-CoV-2 infection. We calculated Pearson correlation coefficients to assess the cross-sectional correlation of immunological parameters at study baseline.

To quantify the associations between immunological parameters and the risk of a SARS-CoV-2 breakthrough infection, we estimated relative risks per 2-fold higher level adjusted for age, sex, and prior SARS-CoV-2 infection. For the majority of parameters (i.e., anti-S IgG, neutralizing antibodies in a sVNT, T-cell response, and anti-N Ig), we analyzed time-to-event data using Cox regression. In these analyses, participants were censored at the time of a SARS-CoV-2 infection, end of the follow-up period, or loss to follow-up, whichever came first. Participants were considered lost to follow-up if they withdrew from the study or had more than one consecutive missing antigen test result, no positive PCR test result, and did not provide anti-N Ig test results at the beginning and the end of the study. The proportional-hazards assumption was tested on the basis of Schoenfeld’s residuals and was met. For the titers of neutralizing antibodies in a pVNT, which was measured only in a nested case-control sample, we used conditional logistic regression to estimate odds ratios for breakthrough infection adjusted for age, sex, and prior SARS-CoV-2 infection. As expected, SARS-CoV-2 events were relatively rare in our study and we therefore describe odds ratios and hazard ratios collectively as measures of relative risk (“rare disease assumption”). We conducted secondary analyses that (i) focused on symptomatic SARS-CoV-2 infections and (ii) quantified relative risks across different categories of baseline antibody levels.

To assess the incremental predictive values of measuring different immunological parameters, we quantified improvements in the C-index when adding them to a model containing information on age and sex.^37^ The C-index is the preferred measure of risk discrimination for time-to-event data and assesses whether the model correctly predicts the order of failure of randomly selected pairs of participants. A C-index of 1.0 indicates perfect prediction of the order of failure; a C-index of 0.5 is achieved purely by chance. P values ≤0.05 were deemed as statistically significant and all statistical tests were two-sided. Analyses were carried out with Stata 15.1 and R 4.1.0.

### Data availability

Data on COVID-19 cases in the district of Schwaz, Tyrol, in Austria is publicly available (data.gv.at - Open Data Österreich). Tabular data on the Shieldvacc-2 cohort can be requested from the corresponding authors by researchers who submit a methodologically sound proposal (including a statistical analysis plan); participant-level data on the Shieldvacc-2 cohort cannot be shared due to regulatory restrictions.

## Data Availability

Data on COVID-19 cases in the district of Schwaz, Tyrol, in Austria is publicly available (data.gv.at - Open Data Oesterreich). Tabular data on the Shieldvacc-2 cohort can be requested from the corresponding authors by researchers who submit a methodologically sound proposal (including a statistical analysis plan); participant-level data on the Shieldvacc-2 cohort cannot be shared due to regulatory restrictions.

## Acknowledgements

We thank Bianca Neurauter, Eva Hochmuth, and Luiza Hoch for excellent assistance when organizing the study and Brigitte Müllauer, Evelyn Peer, Lisa-Maria Raschbichler, Evelyn Peer, and Albert Falch for excellent technical support. We are also grateful to all BMAs in the diagnostic section of Institute of Virology at the Medical University of Innsbruck – especially to Maria Huber. We also thank Sabine Embacher and Kathrin Becker from the Clinical Trial Center of the Medical University of Innsbruck for supporting the coordination of the study.

## Author contributions

PW, DvL, FK, WB, ZB, and JK conceived the study. WB, DvL, SS, and LF designed the questionnaire and the online data collection. ZB, AR, JK, and WB were responsible for laboratory tests. LS, LT, and PW were responsible for the data management. LS and LT performed data cleaning. LS and PW performed the statistical data analysis. HS, BF, TH, DB, MS, CO, DvL, and WB were responsible for on-site data acquisition. DvL, PW, FK, HU, and WB supervised the study. LS, WB, and PW drafted the manuscript. All authors critically reviewed and approved the manuscript.

