## Supplementary Material for "Immune response to 2-dose BNT162b2 vaccination and risk of SARS-CoV-2 breakthrough infection: The Shieldvacc-2 study"

Seekircher, Banki, et al.

### Supplementary methods

Sensitivity and specificity of commercially available serological tests

- **Anti-S IgG:** The manufacturer's recommended cut-off value for positivity of ≥7.1 Binding Antibody Units per milliliter (BAU/mL) has a sensitivity of 99.35% (95% confidence interval [CI]: 96.44-99.97%) at ≥15 days after coronavirus disease 2019 (COVID-19) onset (post-symptom onset), and a specificity of 99.60% (99.22-99.80%).
- **Anti-N Ig:** The manufacturer's recommended cut-off index for positivity of ≥1.0 Cut-off-Index (COI) has a sensitivity of 99.5% (95% CI: 97.0-100.0%) at ≥14 days after COVID-19 onset (post-PCR confirmation), and a specificity of 99.80% (99.69-99.88%).

Surrogate virus neutralization assay

For all participants we evaluated SARS-CoV-2 neutralizing antibodies using the TECO SARS-CoV-2 Neutralization Antibody enzyme-linked immunosorbent assay on the SERION Immunomat (TECOmedical, Sissach, Switzerland) to measure inhibitory effects of neutralizing antibodies blocking the interaction of angiotensin-converting enzyme 2 (ACE2) and receptor binding domain (RBD) of SARS-CoV-2 spike protein. In consultation with the manufacturer, we conducted the analysis in a 1:100 dilution (instead of 1:10) in order to ensure reliable quantification. Following an incubation in a capture-plated pre-coated with ACE2 protein and addition of signal-conjugated RBD, extinction values were measured at 450 nm. Five standard sera of known concentrations in the range of 50 to 5,000 international unit per milliliter (IU/mL) were used to quantify sample antibody levels using a Four Parameter Logistic (4PL) curve fit method. In consultation with the manufacturer the cut-off for positivity was set at ≥200 IU/mL. The manufacturer gives a sensitivity of 99.03% (95% CI: 94.07-99.83%), and a specificity of 100.00% (96.68-100.00%) compared to the plaque reduction neutralization test. Values >5,000 were set to 5,000.

Pseudotype virus neutralization assay

For a subset of 280 participants, we evaluated SARS-CoV-2 neutralizing antibodies using a vesicular stomatitis virus (VSV) pseudovirus-based assay. Briefly, four-fold serial dilutions of heat-inactivated plasma were incubated in duplicates with a replication defective VSV expressing green fluorescent protein (GFP) as marker gene and pseudotyped with a C-terminally truncated version of the spike protein of SARS-CoV-2 (Wuhan isolate) for 1h at 37 °C. Subsequently, plasma/virus mixtures were used to infect subconfluent 293T-ACE2 cells seeded one day before. After approximately 16 hours of infection, the number of GFP positive cells was counted in an ImmunoSpot® S5 analyzer (C.T. L. Europe, Bonn, Germany). 50 % neutralizing antibody titers were calculated via nonlinear regression in GraphPad Prism (version 9) as previously described. Samples with reciprocal neutralization titers >16 were considered positive and ≤16 as negative. Titers >1,024 were set to 1,024.

Interferon-Gamma release assay

In a random sample of 929 participants cellular immune responses were evaluated by QuantiFERON (QFN) SARS-CoV-2 Interferon-Gamma (IFN-γ) release assay (IGRA) (Qiagen, Hilden, Germany). Blood samples were collected in S-Monovette tubes (Sarstedt, Nümbrecht, Germany) containing lithium-heparin anticoagulant (Li-Heparin LH/9mL), stored at room temperature (17-25°C), and processed within ≤8 hours centrally at the laboratory of the Institute of Virology of the Medical University of Innsbruck, Austria. One mL whole blood was loaded onto QFN tubes coated with two spike-derived peptide antigen pools (Ag1 and Ag2) or QFN tubes for a negative control (Nil) and for a positive control (Mitogen). Ag1 contains CD4 whereas Ag2 contains a combination of CD4 and CD8 peptide pools derived from SARS-CoV-2 spike antigen (S1 S2 RDB). After 16-24 hours of incubation at 37°C, tubes were centrifuged for 15 min at 2,000 g, plasma was harvested, and stored at -80°C until used. The IFN-γ concentration in the plasma fraction was measured by QFN human IFN-γ ELISA Kit (Qiagen, Hilden, Germany) according to manufacturer's instructions using an Epoch Microplate Spectrophotometer (BioTek, Vermont, USA). The Qiagen QuantiFERON SARS-CoV-2 IGRA is so far for research use only, no performances and official cut-off value have yet been defined by the manufacturer. Therefore, the ratios of IFN-γ values from SARS-CoV-2 specific stimulation and the unstimulated control was determined as the stimulation index (SI) to mitigate against the background IFN-γ in the sample. We considered samples with SI values $<2$ as negative, $2\leq$ SI $<3$ as weakly reactive, and values $\geq 3$ as reactive.

### Supplementary figures

#### Supplementary Fig. 1: Local and systemic complaints reported within 7 days after first and second dose of SARS-CoV-2 vaccination.

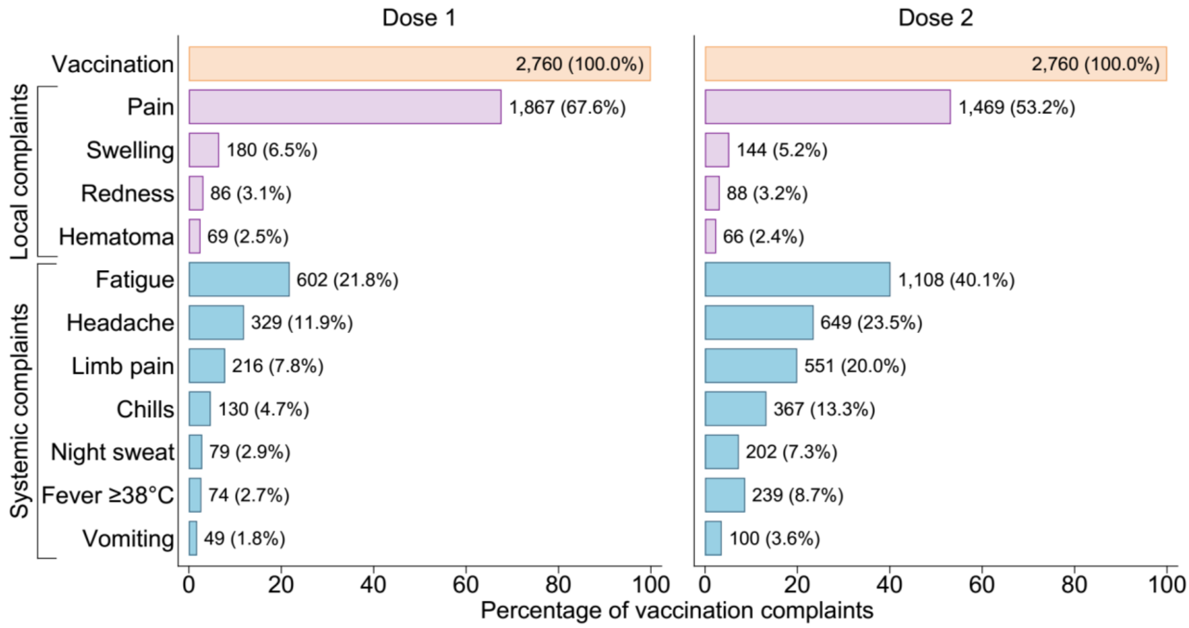


Number and percentage of local and systemic vaccination complaints reported within 7 days after first and second dose of SARS-CoV-2 infection. There were no missing values for vaccination complaints.

### Supplementary Fig. 2: Cumulative incidence of SARS-CoV-2 infection in the Shieldvacc-2 study and in the district of Schwaz

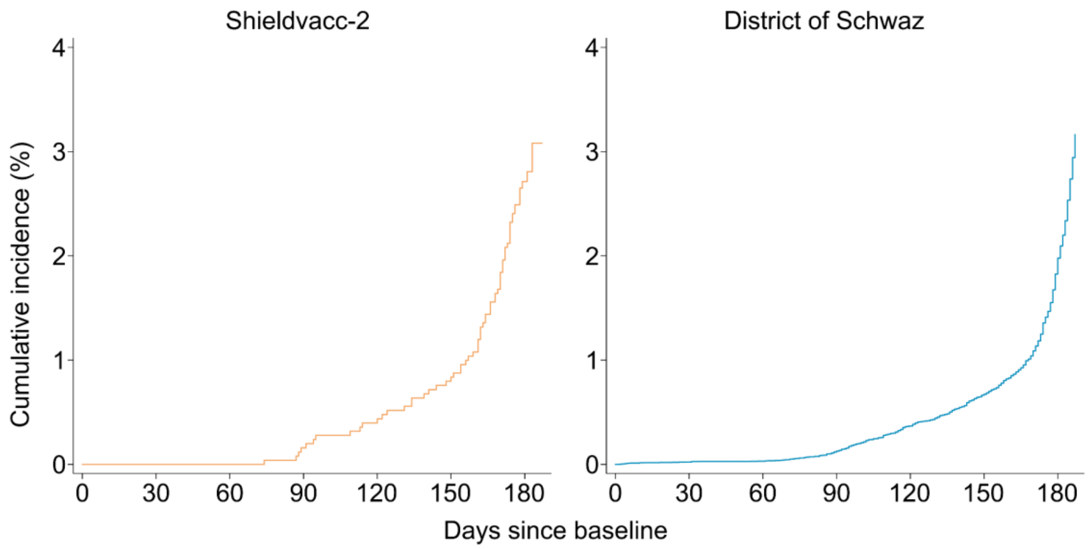


| | | | | | | | |
|---|---|---|---|---|---|---|---|
| No. at risk: | 2,760 | 2,630 | 2,539 | 2,510 | 2,499 | 2,477 | 1,306 |
| No. of events: | 0 | 0 | 4 | 6 | 10 | 46 | 2 |

| | | | | | | | |
|---|---|---|---|---|---|---|---|
| No. at risk: | 84,456 | 84,436 | 84,429 | 84,354 | 84,146 | 83,895 | 82,913 |
| No. of events: | 20 | 7 | 75 | 208 | 251 | 982 | 1,130 |

Kaplan – Meier curves showing cumulative incidence of SARS-CoV-2 infection (%) in the Shieldvacc-2 study and in the district of Schwaz, Tyrol, Austria. Cumulative SARS-CoV-2 incidence of the district of Schwaz was calculated from public data (data.gv.at - Open Data Österreich).

**Supplementary Fig. 3: Symptoms and hospitalization of incident SARS-CoV-2 infections**

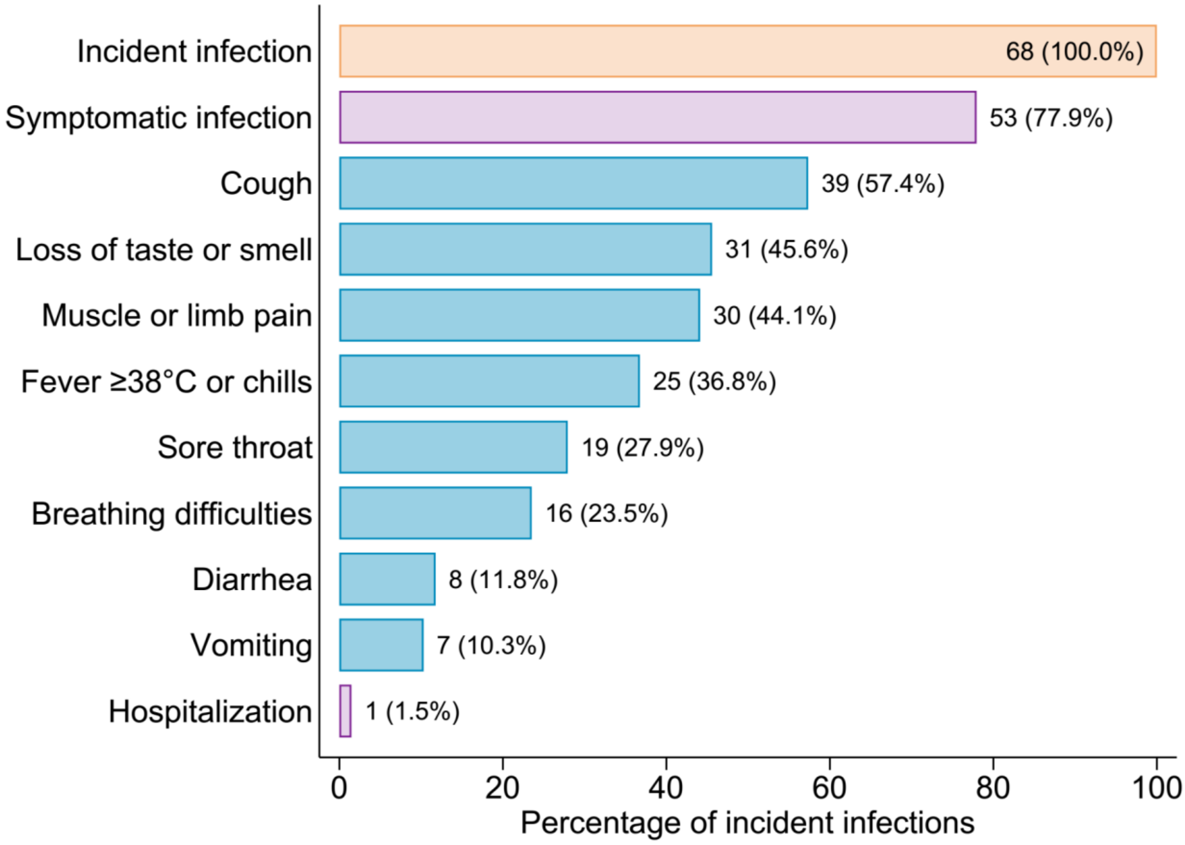


Number and percentage of symptoms and hospitalization during incident SARS-CoV-2 infection.

**Supplementary Fig. 4: Cumulative incidence of SARS-CoV-2 infection in the Shieldvacc-2 study and in the district of Schwaz, Austria**

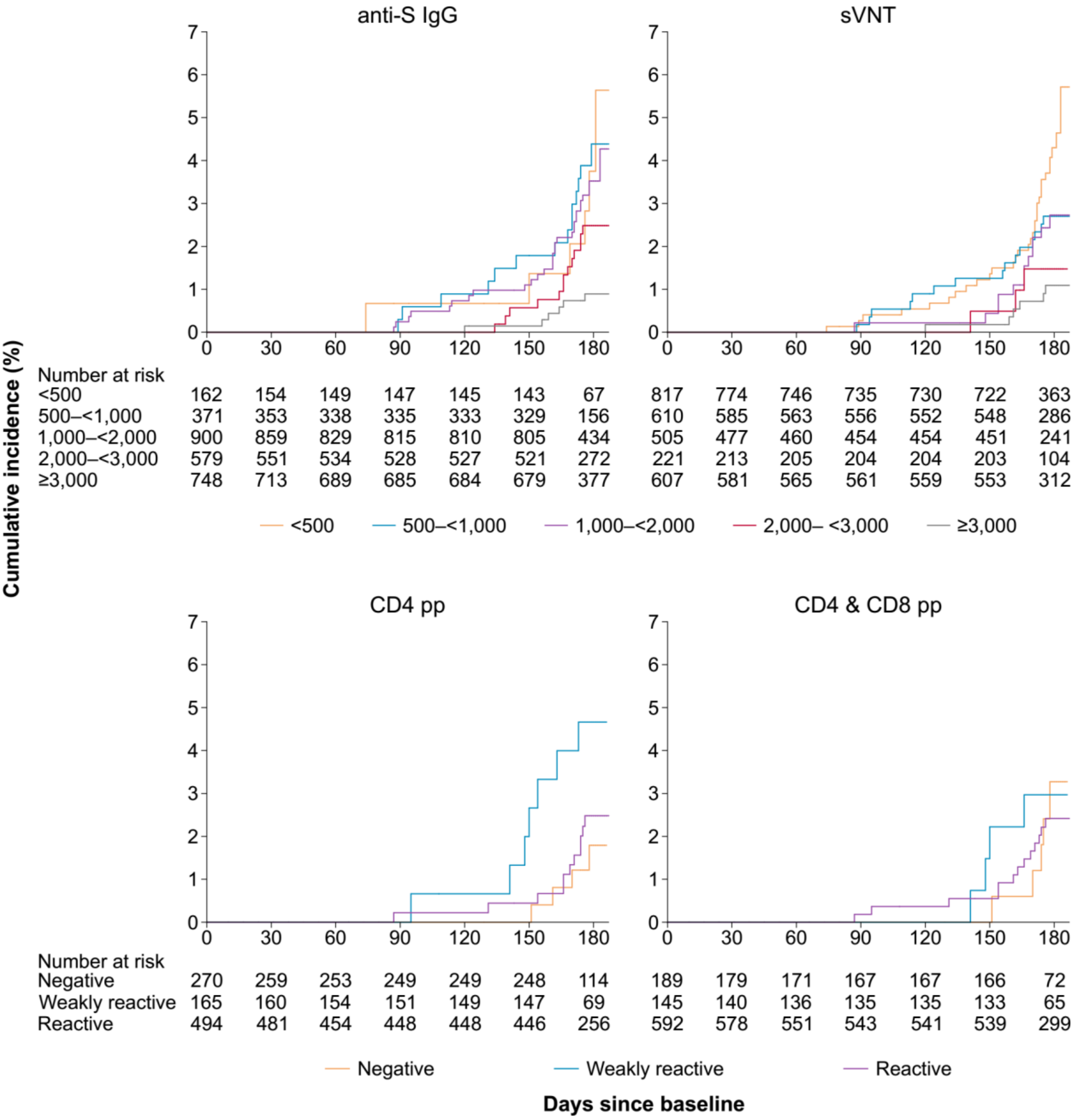


Kaplan – Meier curves showing cumulative incidence of SARS-CoV-2 infection (%) in the Shieldvacc-2 study according to different categories of immunological parameters. Abbreviations: pp, peptide pool; sVNT, surrogate SARS-CoV-2 virus neutralization test.

### Supplementary tables

#### Supplementary Table 1: Information on participants with prior SARS-CoV-2 infection who experienced an incident SARS-CoV-2 infection

| No | Baseline anti-S IgG, BAU/mL | Follow-up anti-S IgG, BAU/mL | Baseline anti-N Ig, COI | Follow-up anti-N Ig, COI | Time since prior infection, months | Symptomatic incident infection* | Method for identifying incident infection |
|---|---|---|---|---|---|---|---|
| 1 | 1,099.9 | 160.9 | 13.07 | 264 | 6.27 | No | Anti-N Ig increase |
| 2 | 779.5 | 130.4 | 2.1 | 10.21 | 6.27 | Yes | Anti-N Ig increase |
| 3 | 3,786.6 | 654 | 1.06 | 4.78 | 14.06 | Yes | Anti-N Ig increase |
| 4 | 2,052.5 | 291.7 | 0.269 | 26.62 | 4.70 | No | Seroconversion |
| 5 | 1,627.3 | 139.2 | 0.278 | 47.11 | 5.85 | No | Seroconversion |
| 6 | 2,717.1 | 453.6 | 47.8 | 216.5 | 4.73 | Yes | PCR |
| 7 | 1,887.9 | 263.3 | 4.64 | 116.7 | 6.57 | No | Anti-N Ig increase |
| 8 | 50.9 | 14.4 | 0.062 | 2.4 | 6.57 | No | Seroconversion |
| 9 | 1,321.6 | 3,890.9 | 3.14 | 15.29 | NA | Yes | PCR |

*Symptomatic SARS-CoV-2 infection was defined as having one or more symptoms including fever or chills, cough, breathing difficulties, muscle or limb pain, lost sense of smell or taste, sore throat, diarrhea, or vomiting. Abbreviations: BAU, Binding Antibody Units; COI, Cut-off-Index; mL, milliliter; NA, not available.
